# Trajectory of the body mass index of children and adolescents attending a reference mental health center

**DOI:** 10.1101/2024.01.12.24301227

**Authors:** Juliana Echeveste-Navarrete, Patricia Zavaleta-Ramírez, Maria Fernanda Castilla-Peon

## Abstract

**Objectives:** To describe the standardized body mass index (z-BMI) trajectory of children and adolescents admitted to a psychiatric reference center in Mexico City according to their diagnosis and medication use. A secondary objective was to compare z-BMI between antipsychotic users and non-users.

**Methods:** Diagnosis, prescribed medications, serial heights, and weights were collected from the medical records.

**Results:** The median baseline z-BMI of the 129 analyzed cases was 0.88 (IQR: 0 -1.92), and the overweight/obesity prevalence was 46.8%. At the end of follow-up (median, 50.3 weeks), they had a median change in z-BMI of -0.09 (IQR: - 0.68 -0.42). New long-term users of antipsychotics (n = 29) had an increase in their z-BMI, in contrast to never-users (median difference 0.73, p = 0.01) and to previous users (median difference 0.92, p = 0.047). The 59 subjects with excessive weight at admission had a median change in z -BMI of -0.39 (IQR: - 0.81--0.04). Among patients with excessive weight and depression, a greater decrease in z-BMI was observed in sertraline users (n = 13) compared with fluoxetine users (n = 15) (median -0.65 vs. 0.21, p<0.001).

**Conclusions:** New long-term users of antipsychotics increased their z-BMI significantly. Patients with depressive disorders and obesity on sertraline therapy tend to decrease their z-BMI.

## Introduction

Obesity is one of the most prevalent and measurable cardiovascular risk factors with the potential to be modified. Mexico has the highest prevalence of pediatric obesity in the world, with 38% of children 5 to 11 years old and 43% of adolescents 12 to 19 having overweight or obesity in Mexico City. (1) On the other hand, mortality from cardiovascular and cerebrovascular diseases in people with psychiatric conditions is 50 to 90% higher than in the general population. (2)

Two groups of patients with psychiatric conditions are of particular interest. First, patients on antipsychotics have metabolic disturbances such as weight gain and lipid alterations, with variability among different drugs and a possible independent effect of their underlying conditions. (3,4) The second group of interest is children and adolescents with depressive disorders. Depression per se is a known cardiovascular risk factor. (5–7) Other cardiovascular risk markers are associated with depression, obesity being one of the most studied. Children with obesity have higher rates of mental disorders compared with normal-weighted children. (8) Moreover, longitudinal studies in adolescents have shown that depression in adolescence is a predictor of elevated body mass index (BMI) in adulthood (9), and being overweight in childhood increases the odds of having a lifetime mood disorder (10). As a population with a high prevalence of cardiometabolic risk factors, it is mandatory to estimate the metabolic secondary effects of antipsychotics in Mexican children and adolescents.

The primary aim of this study was to describe the trajectories of standardized BMI (z-BMI) in a cohort of pediatric users of a mental health tertiary-care center in Mexico City as a group and by clinical subpopulations. An exploratory aim was to find an association between z-BMI and the prescribed drugs. We hypothesized that patients receiving antipsychotic medication would increase their BMI more than patients without antipsychotics.

## Materials and Methods

This was a retrospective cohort study with data collected from medical records. We included every subject admitted to the Psychiatric Children Hospital ‘Dr. Juan N. Navarro’ from January 1st, 2022, both ambulatory and as an inpatient, if they had a follow-up of at least six months. Consecutive medical files were checked until the sample size was reached on March 25, 2022.

We extracted demographic data and the psychiatric diagnoses registered at their initial in-depth interview, according to the International Classification of Diseases, version 10th (ICD-10) codes. We also extracted information on all medications prescribed and serial data on body weight and height during subsequent visits with their psychiatrist.

Statistical analysis was performed with Stata/MP, version 14.0 (StataCorp, College Station, TX, USA). We calculated the BMI z-score (z-BMI) for age and sex according to the World Health Organization standards with the STATA WHO 2007 package (De Onis et al., 2007). Excessive weight was defined as a z-BMI >1, obesity as a z-BMI >2, and overweight as a z-BMI between 1 and 2. Anthropometric data for cases with a |z-BMI| >4 were verified in clinical records, and unplausible heights were treated as missing data (n=7). We collected data from each subject at a maximum of five time points. Since periodicity in anthropometric measures was highly variable, we analyzed timing as follows: the first visit (t0) was the initial assessment at admission; the second visit (t1) at a median of 9.3 weeks (range: 6-12); the third visit (t2) at a median of 17.7 weeks (range: 12.1 - 23), fourth visit (t3) at 30.3 weeks (range: 23.1 - 39) and the fifth visit (t4) at 50.3 weeks (range: 42-57). ICE-10 codes F32 and F33 were grouped as “Depressive disorders,” F40-F48 were grouped as “Anxiety disorders,” F50 as “Eating disorders,” F84 as “developmental disorders, and “F90” as Attention-deficit hyperactivity disorders. We grouped antipsychotic users into four groups: previous users, never users, short-term new users (had an antipsychotic prescription in 1-3 time points), and long-term new users (had an antipsychotic medication in at least four time points).

Descriptive statistics are reported as medians and interquartile ranges (IQR) for quantitative variables and frequencies for qualitative ones. Statistical significance for median comparison was performed with Mann-Whitney U-test, paired samples, Wilcoxon matched-pairs signed-rank test, or Friedmańs test as appropriate. The Fisher exact test and binomial distribution were used for frequency comparisons. Statistical significance was set at a two-tailed α of 0.05. The sample size was calculated with an 80% power to find a difference of 0.5 z-BMI units between groups (i.e., with or without antipsychotics), assuming a standard deviation of 1.

This study has complied with the World Medical Association Declaration of Helsinki regarding ethical conduct of research involving human subjects. The Ethics and Research Board of the Psychiatric Children Hospital’s ‘Dr. Juan N. Navarr0’ approved the study with the registration number OI1/02/1122 on February 18^th^, 2023.

## Results

From January 1, 2022, to March 24, 2022, there were 492 new files, from which 129 met the follow-up of at least six months criterion to be included. The median age at the first visit was 11.9 years (IQR: 7.3-14.2), and 70 (54.7%) were male. The most frequent diagnosis was a depressive disorder in 68 cases (52.7%), followed by anxiety disorders in 49 patients (38%) and developmental disorders in 30 cases (23.2%). The median follow-up was 50.3 weeks, with a minimum of 37.3 and a maximum of 66.3. During follow-up, 99 (77%) of cases had at least five measurements of body weight, and 26 (20%) had four. Further basal description of diagnoses and medications is presented in Table 1.

**Table 1.** Baseline characteristics of the study sample.

|  | All | Depressive disorder | Developmental Disorder | Anxiety disorder | Antipsychotics on the first visit | Age 5-11 years | Age 12-19 years |
| --- | --- | --- | --- | --- | --- | --- | --- |
|  | n=129 | n=68 | n=30 | n=49 | n=40 | n=48 | n= 63 |
| **Age (years)** | 11.9 (7.3-14.2) | 13.7 (12.2-15.2) | 4.4 (3.4-7.3) | 13.9 (12.7-15.2) | 13.6 (10.2-14.6) | 9.2 (6.8-10.3) | 14.2 (13.3-15.5) |
| **Male sex (n,%)** | 70 (54.7%) | 23 (34.3%) | 28 (93.3%) | 11 (22.9%) | 23 (57.5%) | 32 (66.7%) | 20 (32.6%) |
| **Depressive disorder** | 68 (52.7%) | - | 0 | 40 (81.6%) | 23 (57.5%) | 15 (31.3%) | 53 (84.1%) |
| **Developmental disorder** | 30 (23.3%) | 0 | - | 0 | 6 (15%) | 11 (22.9%) | 2 (3.17%) |
| **Anxiety disorder** | 49 (38%) | 24 (35.3%) | 0 | - | 8 (20%) | 9 (18.7%) | 40 (63.5%) |
| **Eating disorder** | 8 (6.2%) | 7 (10.3%) | 0 | 5 (10.2%) | 4 (10.0%) | 1 (2.1%) | 7 (11.1%) |
| **Any antipsychotic** | 40 (31.1%) | 23 (33.8%) | 6 (20%) | 14 (35%) | - | 11 (22.9%) | 27 (42.9%) |
| **Risperidone** | 26 (20.2%) | 15/68 (22.1%) | 4/30 (13.3%) | 8/49 (16.3%) | 26/40 (65%) | 9 (18.7%) | 15 (23.8%) |
| **Quetiapine** | 8 (6.2%) | 5/68 (7.4%) | 0 | 5 (62.5%) | 8/40 (20%) | 1 (2%) | 7 (11.1%) |
| **Olanzapine** | 4 (3.1%) | 0 | 0 | 2 (4.1%) | 4/40 (10%) | 0 | 4 (6.4%) |
| **Haloperidol** | 2 (1.55) | 0 | 1 (3.3%) | 0 | 2/40 (5%) | 0 | 2 (3.2%) |
| **Methylphenidate /atomoxetine any time** | 30 (23.3%) | 12 (17.7%) | 6 (20%) | 5 (10.2%) | 11 (27.5%) | 17 (56.7%) | 10 (33.3%) |

At their first visit, the median z-BMI was 0.88 (IQR: 0-1.92) for the whole sample, with a prevalence of obesity and excessive weight of 23% and 46.8%, respectively. These frequencies are higher than those reported for Mexico Citýs general pediatric population of 15% (p = 0.006) and 40% (p=0.07) (Table 2, Figure 1). (1)

**Table 2.**
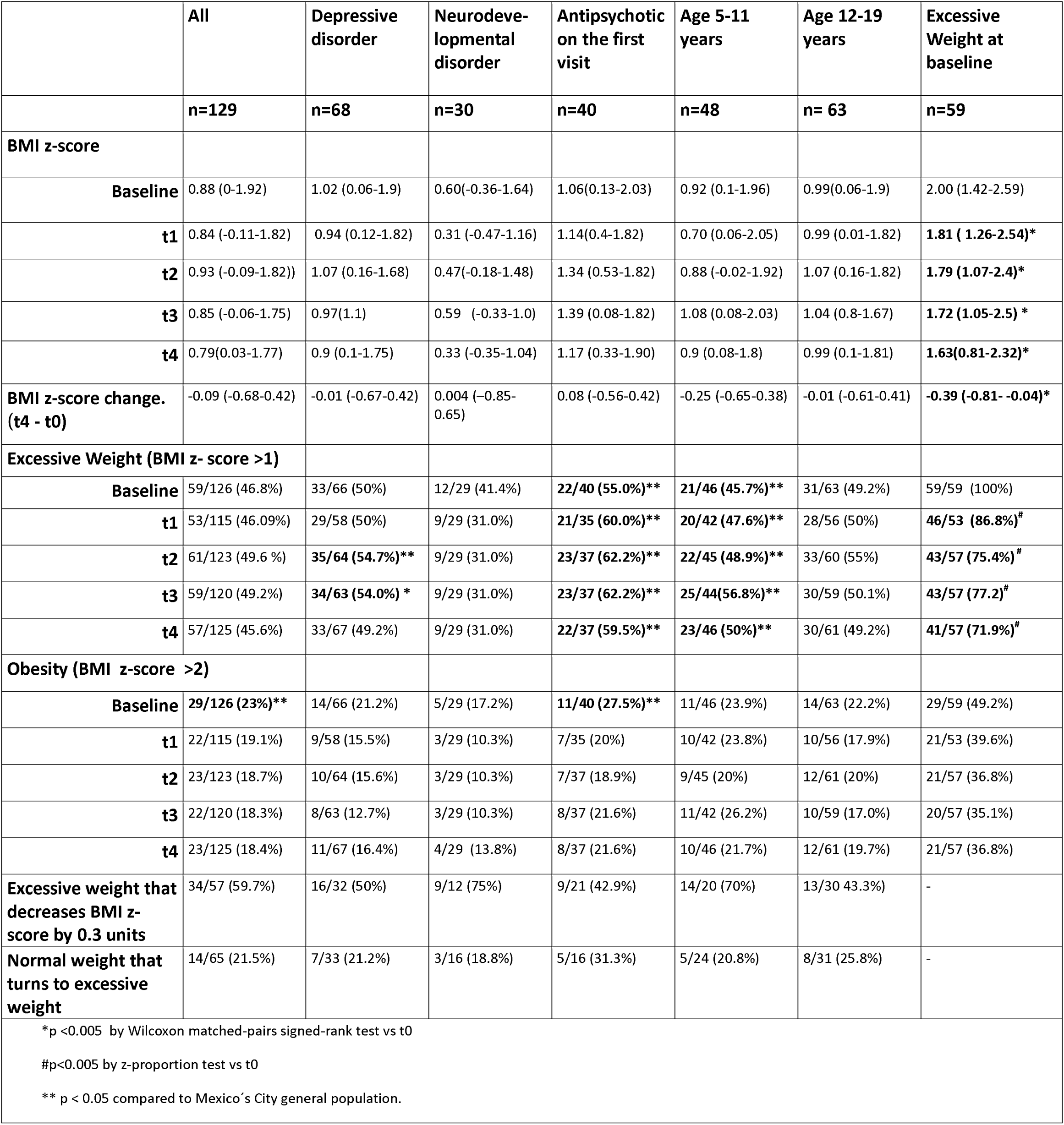
Anthropometric characteristics at follow-up.

**Figure 1.**
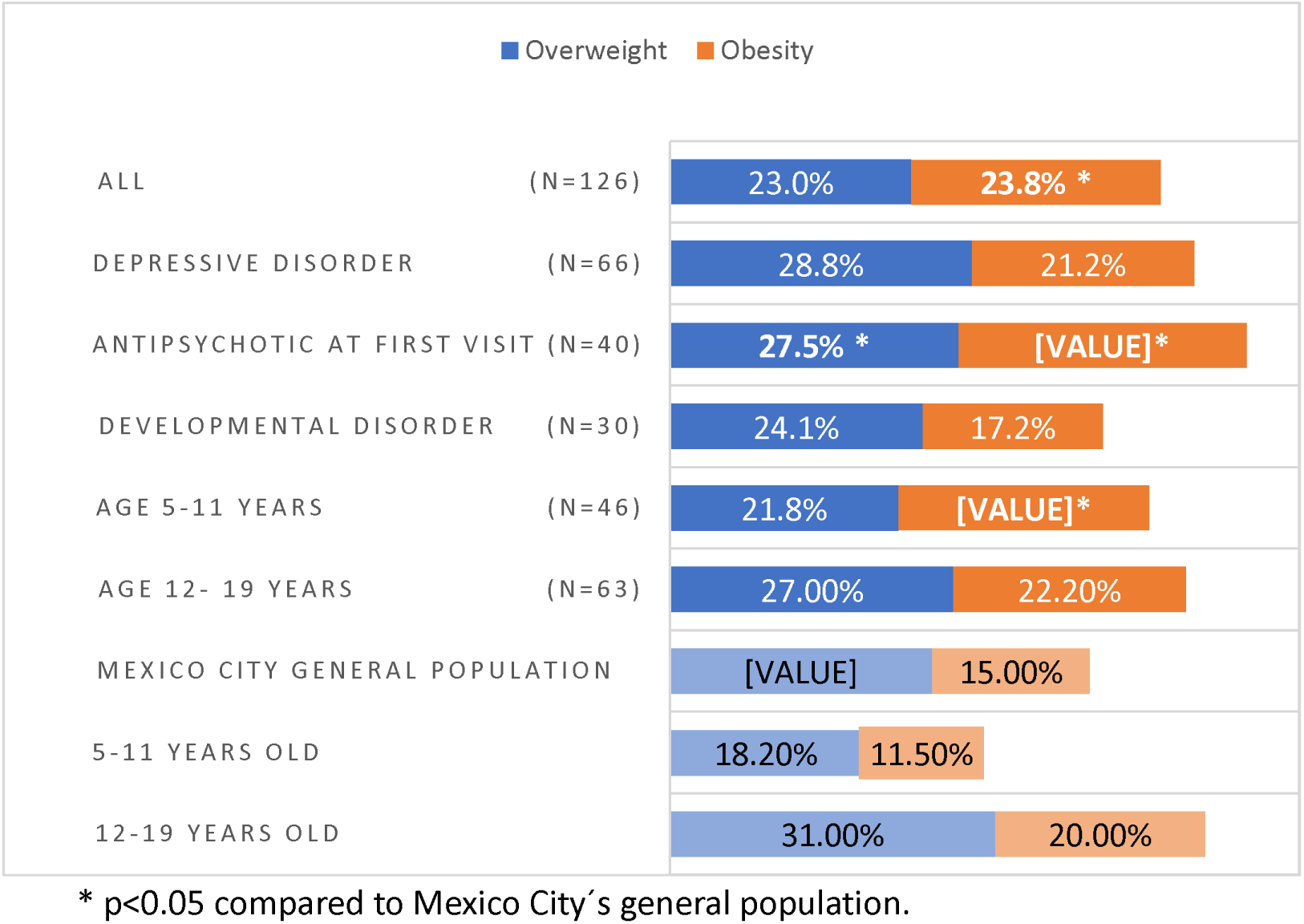
Prevalence of overweight and obesity at baseline.

### Patients on antipsychotics

Regarding antipsychotics, 73 cases (57%) received one of these drugs in at least one visit. Fourteen (10.9%) started them before their admission, 40 (31%) had one of these medications prescribed on their first visit, and 32 (24.8%) were long-term new users. Depressive disorder was the most frequent diagnosis in patients who were prescribed an antipsychotic on their first visit (57.5%). The most frequently prescribed antipsychotic was risperidone (65%), followed by quetiapine (20%), olanzapine (10%), and haloperidol (2. 5%).

Median z-BMI (1.06, IQR: 0.13-2.03), excessive weight (55%), and obesity (27.5%) were higher in patients who had antipsychotics prescribed on their first visit, without statistical significance, compared to cases without antipsychotics. New long-term users of antipsychotics (n = 29) significantly increased their z-BMI compared to previous users (median difference=0.92, p= 0.047) and to never users (median difference = 0.70, p = 0.01). **(Figure 2)**

**Figure 2.**
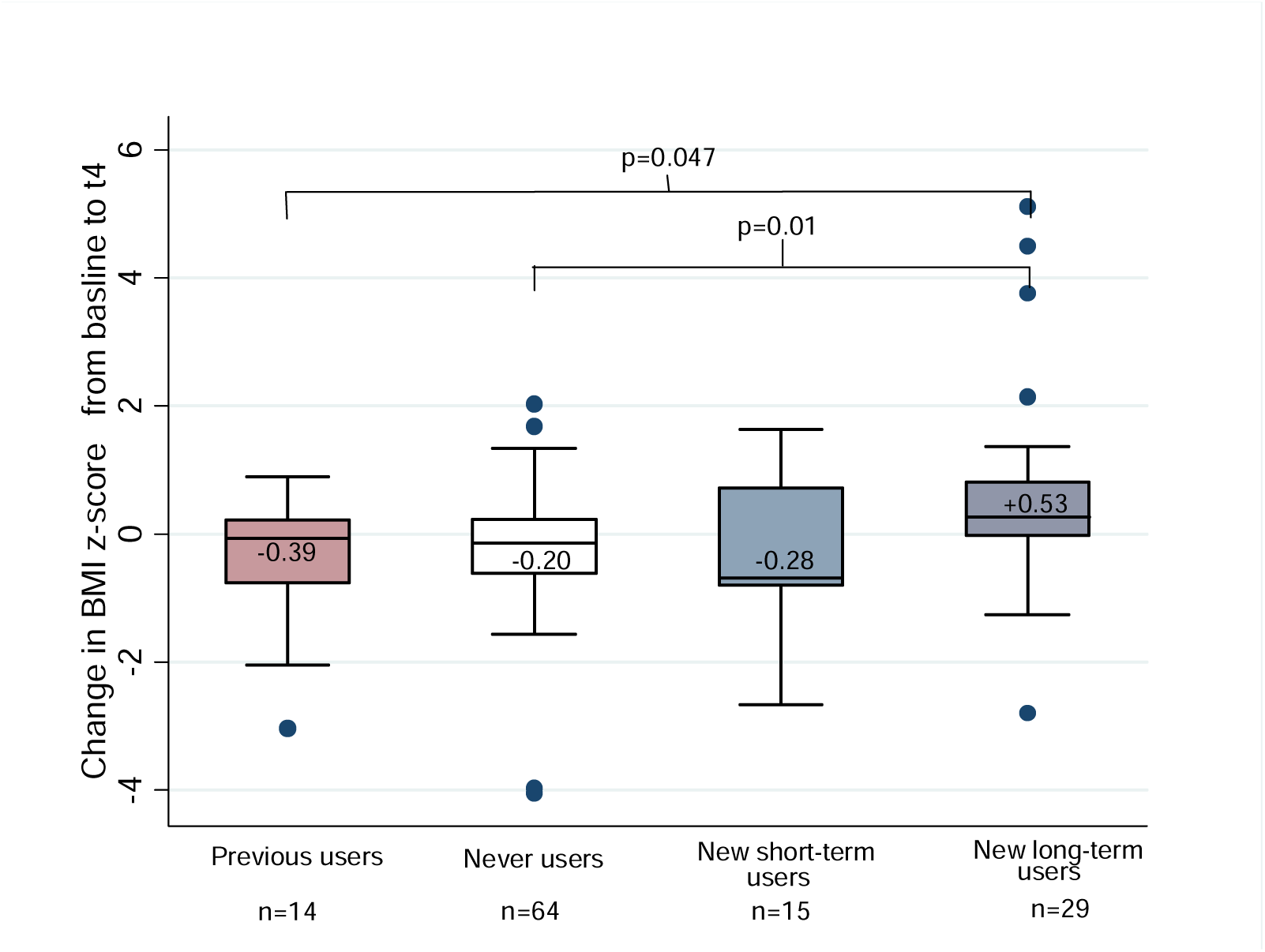
Change in z-BMI according to antipsychotic use.

### Patients with depression

Sixty-eight patients had depression on their first visit, and 24 of them (35.3%) had an anxiety disorder as a comorbidity. This group had a median z-BMI of 1.02 (IQR: 0.06-1.9) without a significant change in the follow-up, and thirty-three (50%) had excessive weight at baseline. However, cases with both depression and excessive weight (n = 32) at baseline did have a median decrease in their z-BMI of 0.36 units (IQR: -0.76-0.12, p =0.008) **(Figure 3)**. Patients with depression and excessive weight on sertraline (n=13) significantly decreased their median z-BMI (-0.63, p = 0.003) while fluoxetine users (n = 15) did not (+0.21, p = 0.55). **(Figure 4)** The difference between median z-BMI change at the end of follow-up between patients with excessive weight treated with sertraline vs. fluoxetine was 0.55 (p = 0.01). Fluoxetine users had a higher prevalence of antipsychotic use than sertraline users, but the difference in z-BMI change did not change when adjusted for this variable (Table 3).

**Figure 3.**
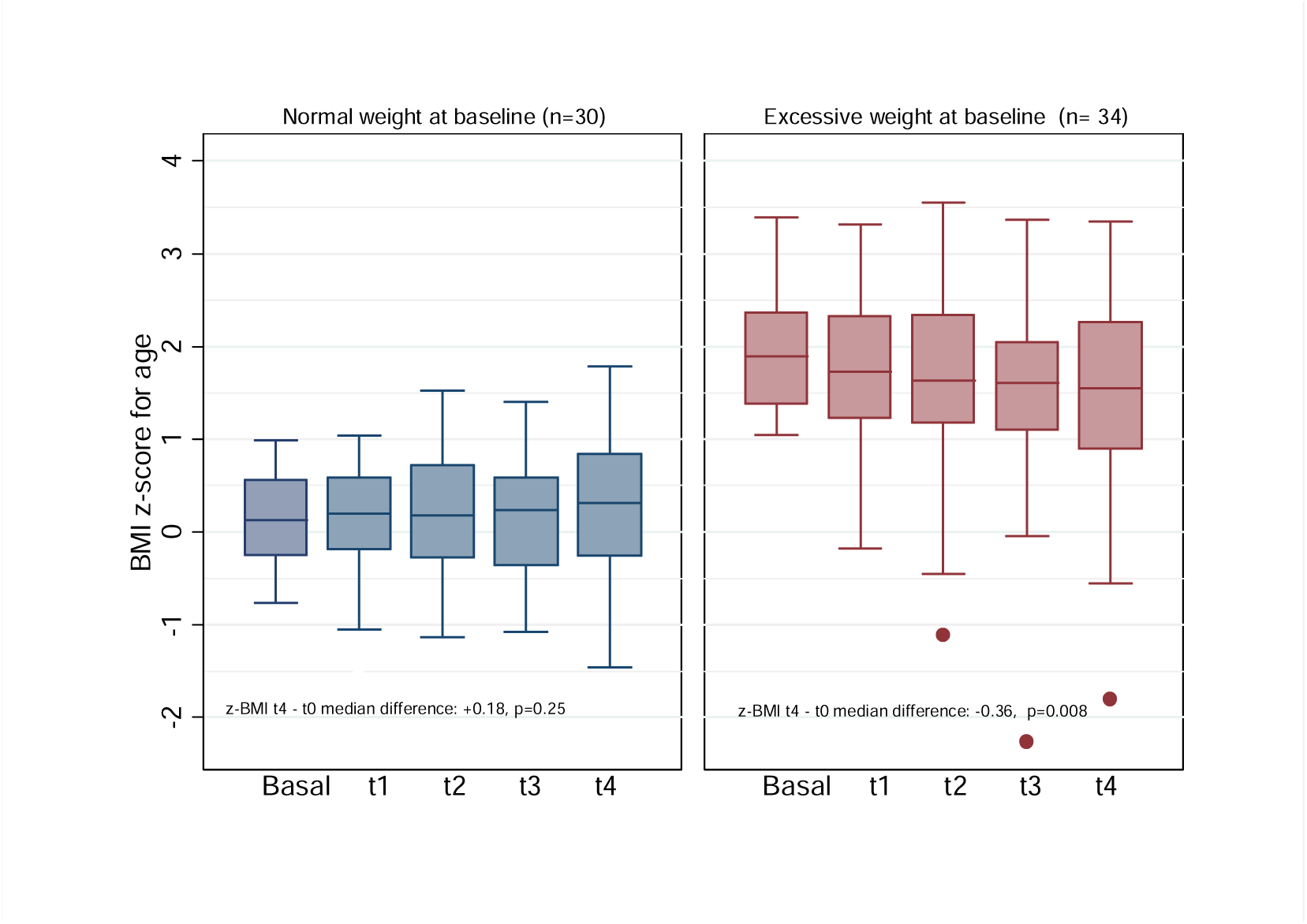
BMI-z score trajectory in cases with a depressive disorder at baseline.

**Figure 4.**
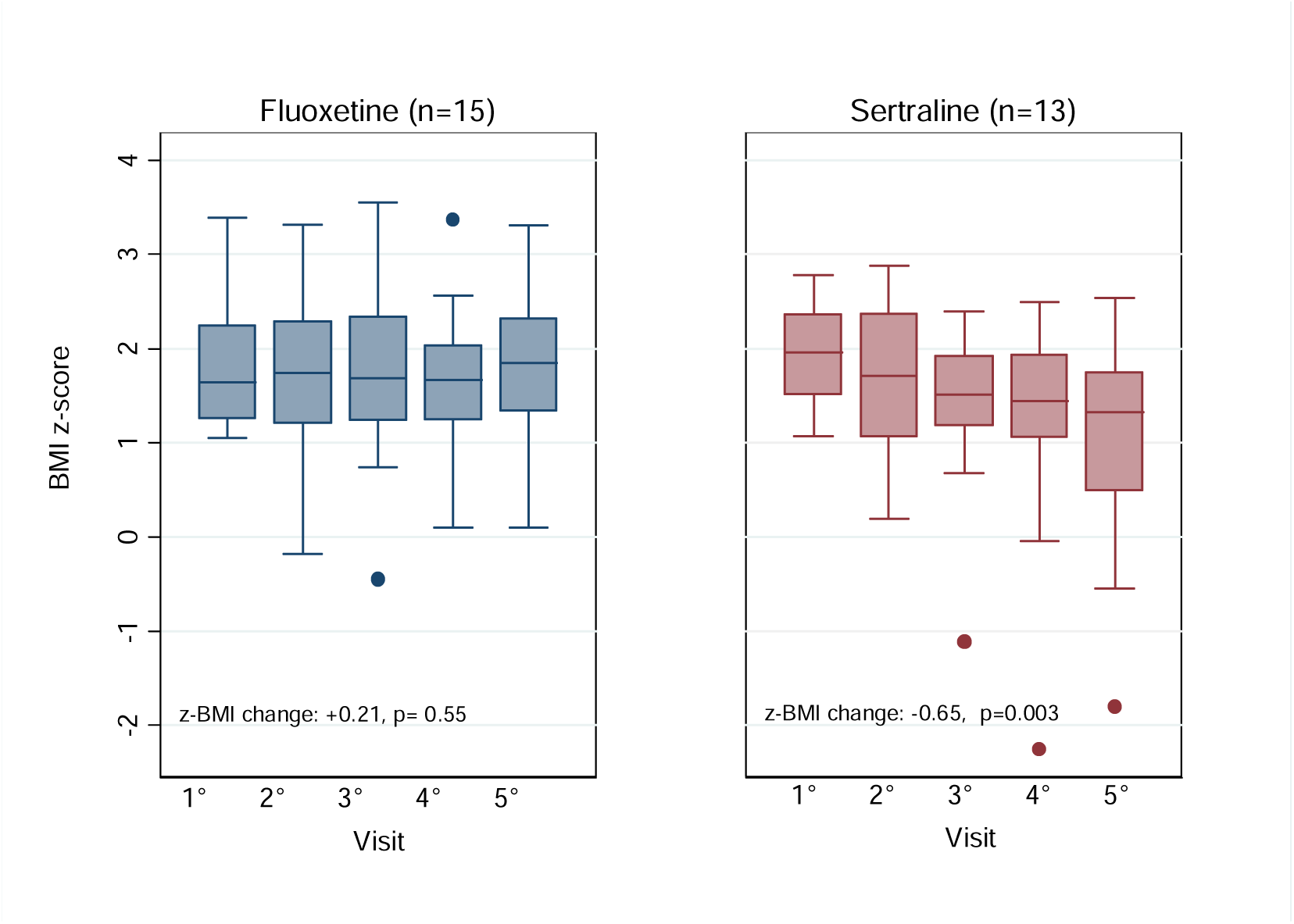
BMI z-score trajectory in cases with depression and excessive weight by antidepressive used.

**Table 3.** Cases with a depressive disorder and excessive weight.

**Tab 3. Cases with a depressive disorder and excessive weight.**
|  | **Fluoxetine**<br>**N=15** | **Sertraline**<br>**N=13** | **p** |
| --- | --- | --- | --- |
| **Age (median, IQR)** | 13.8 (11.5-15.2) | 12.3 (11.9-13.4) | 0.32 |
| **Male, n (%)** | 3 (20.3%) | 4 (30.8 %) | 0.78 |
| **BMI z-score at baseline** | 1.64 (1.26-2.25) | 1.96 (1.52-2.36) | 0.46 |
| **BMI z-score change t4-t0 (median, IQR)** | -0.05 (-0.31 -0.26) | -0.69 (-0.81-0.39) | **0.01** |
| **Obesity at baseline, n (%)** | 6 (40%) | 6 (46.2%) | 0.52 |
| **Obesity at t4, n (%)** | 6 (42.9%) | 3 (23.1%) | 0.25 |
| **Anxiety disorder, n (%)** | 7 (46.7%) | 8(61.5%) | 0.48 |
| **Eating disorder, n (%)** | 1 (14.3%) | 1 (14.3%) | 1.00 |
| **Risperidone any time, n(%)** | **10 (66.7%)** | **3 (23.1 %)** | **0.03** |
| **Quetiapine any time, n(%)** | 5 (33.3%) | 2 (15.2%) | 0.4 |
| **Antipsychotic any time , n(%)** | **11 (73.3 %)** | **5 (28.5%)** | **0.05** |
| **Stimulant any time**<br>**(Methylphenidate / Atomoxetine) n(%)** | 2 (13.37%) | 2 (15.4%) | 1.00 |

## Discussion

To our knowledge, this study is the first to describe weight status trajectories among Mexican pediatric patients with mental disorders. We found a baseline prevalence of excessive weight of 46.8 % and 23.8% of obesity in our sample, higher than the reported in Mexico Citýs children and adolescent general population. (1) Excessive weight was especially prevalent in antipsychotic users and patients diagnosed with a depressive disorder. Other studies in children treated with second-generation antipsychotics have shown a prevalence of obesity of 31% and overweight of 26%, which is similar to our results. (11) As a group, school-aged children had a significantly higher proportion of obesity compared to the general population.

Genetic and lifestyle conditions shared with the general Mexican population may partly explain our sample’s high prevalence of excessive weight. Additionally, obesity has been associated with mental disorders such as mood disorders, attention deficit hyperactivity disorder, binge eating, and schizophrenia. (12) Involved mechanisms may include increased unhealthy behaviors in this population, medication side effects, and possibly shared molecular and immunological pathways. (2)

Regarding trajectories, we found a significant increase in z-BMI among new long-term users of antipsychotics. Cases who were on antipsychotics before admission had a high z-BMI at baseline but did not show a significant further increase. This is consistent with most reports, which show a higher weight gain during the first months of treatment with antipsychotics with a later stabilization. (13,14) Weight gain in antipsychotic-naïve children has been reported .in 84.4% of patients on olanzapine, 64% on risperidone, and 55.6% on quetiapine. (15,16) In our sample, the most frequently prescribed antipsychotic was risperidone, which is indicated to manage psychotic symptoms and modulate aggressive behavior in children and adolescents with neurodevelopmental and/or conduct disorders. (17,18)

Drug-induced disturbances in hormones and neurotransmitters involved in the hypothalamic regulation of food intake and energy metabolism (i.e., leptin, adiponectin, ghrelin, dopamine, histamine, and serotonin) may contribute to antipsychotic-induced weight gain (14,16).

Regarding depression and obesity, plenty of evidence in the adult population indicates a bidirectional association between these two conditions. (19–21) However, longitudinal studies in the pediatric population report inconsistent results about the causal role of obesity on depression and insufficient exploration of depression as a risk factor for obesity. (22–26)

An interesting finding in our study was an unexpected and important reduction in z-BMI in people with depression and excessive weight at baseline. The size effect observed in this study (-0.36 standard deviations) is about three times the expected from intensive nutritional interventions in adolescents with obesity. (26,27) Remarkably, this population received no specialized nutritional intervention besides standard general pediatric counseling. As we did, some other studies have found an association between an improvement in depression and a reduction in obesity. (28) In a longitudinal intervention study in adolescents with depression, Bai et al. reported a bidirectional association between health risk behaviors and severe depression. However, the weight category did not change over time. (29) A study by Mansoor et al. on adolescents with treatment-resistant depression did not find an association between successful treatment of depression and z-BMI. However, his study did not separately analyze the group with excessive weight at baseline. (30)

In an exploratory analysis, we found a greater reduction in z-BMI in patients with depression and obesity treated with sertraline than in those who received fluoxetine, which is inconsistent with general knowledge. (31,32) Although multivariate analysis suggests that the difference in weight gain persists after adjustment for antipsychotic use, which was more frequent in the fluoxetine group, prospective studies with larger sample sizes and more detailed clinical data should be conducted to conclude.

Our study has some limitations, mainly because of the secondary data source of the information, which had some inconsistencies and missing data. Besides, the sample size is insufficient to analyze confusion and interaction among factors related to weight gain. Finally, crucial cardiometabolic risk factors other than weight status could not be evaluated because of a lack of data.

This exploratory analysis poses some critical clinical questions that might be approached with future prospective studies designed for this purpose. Among these unanswered questions is the role of treatment of childhood depression and anxiety in the improvement of cardiometabolic risk factors, the optimal follow-up of pediatric users of antipsychotics, and the potential role of interventions (i.e., metformin) to prevent weight gain in this setting.

## Conclusion

New long-term users of antipsychotics increased their z-BMI significantly, so it is necessary to emphasize psychoeducation on healthy lifestyles at the beginning of treatment with these medications. More research is needed on the effects of depression and anxiety and their treatment on cardiometabolic risk factors in the pediatric population.

## Declaration of Interest

The authors report that there are no competing interests to declare.

## Data Availability Statement

Data are available on request from the authors to the corresponding author (MFCP).

## Notes

### Competing Interest Statement

The authors have declared no competing interest.

### Funding Statement

This study received no funding

### Author Declarations

Hospital Psiquiatrico Infantil Dr. Juan N. Navarro. Comision Nacional de Salud Mental y Adicciones. The Ethics Review Board approved this study with the registration number OI1/02/1122 on February 18th, 2023.

## References

1. Instituto Nacional de Salud Pública. Datos abiertos de México-INSP-Instituciones [Internet]. [cited 2023 May 17]. Available from: datos.gob.mx/busca/dataset/encuesta-nacional-de-salud-y-nutrición-continua-covid-19-2020-modulo-nutricion

2. Chao AM, Wadden TA, Berkowitz RI. Obesity in Adolescents with Psychiatric Disorders. Vol. 21, Current Psychiatry Reports. Current Medicine Group LLC 1; 2019.

3. Ringen PA, Engh JA, Birkenaes AB, Dieset I, Andreassen OA. Increased mortality in schizophrenia due to cardiovascular disease - a non-systematic review of epidemiology, possible causes and interventions. Front Psychiatry. 2014;5(SEP).

4. Bernardo M, Rico-Villademoros F, García-Rizo C, Rojo R, Gómez-Huelgas R. Real-World Data on the Adverse Metabolic Effects of Second-Generation Antipsychotics and Their Potential Determinants in Adult Patients: A Systematic Review of Population-Based Studies. Adv Ther. 2021 May 138(5):2491–512.

5. Virani SS, Newby LK, Arnold S V., Bittner V, Brewer LPC, Demeter SH, et al. 2023 AHA/ACC/ACCP/ASPC/NLA/PCNA Guideline for the Management of Patients With Chronic Coronary Disease: A Report of the American Heart Association/American College of Cardiology Joint Committee on Clinical Practice Guidelines. Vol. 148, Circulation. NLM (Medline); 2023. p. e9–119.

6. Sumner JA, Khodneva Y, Muntner P, Redmond N, Lewis MW, Davidson KW, et al. Effects of concurrent depressive symptoms and perceived stress on cardiovascular risk in low- and high-income participants: Findings from the Reasons for Geographical and Racial Differences in Stroke (REGARDS) study. J Am Heart Assoc. 2016 Oct 1;5(10).

7. Lichtman JH, Bigger JT, Blumenthal JA, Frasure-Smith N, Kaufmann PG, Lespérance F, et al. Depression and coronary heart disease: Recommendations for screening, referral, and treatment - A science advisory from the American Heart Association Prevention Committee of the Council on Cardiovascular Nursing, Council on Clinical Cardiology, Council on Epidemiology and Prevention, and Interdisciplinary Council on Quality of Care and Outcomes Research. Vol. 118, Circulation. 2008. p. 1768–75.

8. Halfon N, Larson K, Slusser W. Associations between obesity and comorbid mental health, developmental, and physical health conditions in a nationally representative sample of us children aged 10 to 17. Acad Pediatr. 2013 Jan;13(1):6–13.

9. Soczynska JK, Kennedy SH, Woldeyohannes HO, Liauw SS, Alsuwaidan M, Yim CY, et al. Mood disorders and obesity: Understanding inflammation as a pathophysiological nexus. Vol. 13, NeuroMolecular Medicine. 2011. p. 93–116.

10. Gibson-Smith D, Halldorsson TI, Bot M, Brouwer IA, Visser M, Thorsdottir I, et al. Childhood overweight and obesity and the risk of depression across the lifespan. BMC Pediatr. 2020 Jan 21;20(1).

11. Panagiotopoulos C, Ronsley R, Davidson J. Increased Prevalence of Obesity and Glucose Intolerance in Youth Treated With Second-Generation Antipsychotic Medications. Vol. 54, The Canadian Journal of Psychiatry. 2009.

12. Avila C, Holloway AC, Hahn MK, Morrison KM, Restivo M, Anglin R, et al. An Overview of Links Between Obesity and Mental Health. Vol. 4, Current obesity reports. 2015. p. 303–10.

13. Goltz J, Ivanov I, Rice TR. Second generation antipsychotic-induced weight gain in youth with autism spectrum disorders: a brief review of mechanisms, monitoring practices, and indicated treatments. Vol. 67, International Journal of Developmental Disabilities. Taylor and Francis Ltd.; 2021. p. 159–67.

14. Reynolds GP, McGowan OO. Mechanisms underlying metabolic disturbances associated with psychosis and antipsychotic drug treatment. Vol. 31, Journal of Psychopharmacology. SAGE Publications Ltd; 2017. p. 1430–6.

15. Correll CU, Manu P, Olshanskiy V, Napolitano B, Kane JM, Malhotra AK. Cardiometabolic risk of second-generation antipsychotic medications during first-time use in children and adolescents. JAMA. 2009;302(16):1765–73.

16. Krause M, Zhu Y, Huhn M, Schneider-Thoma J, Bighelli I, Chaimani A, et al. Efficacy, acceptability, and tolerability of antipsychotics in children and adolescents with schizophrenia: A network meta-analysis. Vol. 28, European Neuropsychopharmacology. Elsevier B.V.; 2018. p. 659–74.

17. Fitzpatrick SE, Srivorakiat L, Wink LK, Pedapati E V., Erickson CA. Aggression in autism spectrum disorder: Presentation and treatment options. Vol. 12, Neuropsychiatric Disease and Treatment. Dove Medical Press Ltd; 2016. p. 1525–38.

18. Pringsheim T, Hirsch L, Gardner D, Gorman D. The Pharmacological Management of Oppositional Behaviour, Conduct Problems, and Aggression in Children and Adolescents With Attention-Deficit Hyperactivity Disorder, Oppositional Defiant Disorder, and Conduct Disorder: A systematic Review and Meta-Analysis. Part 2: Antipsychotics and Traditional Mood Stabilizers. Canadian Journal of Psychiatry. 2015;60(2):5261.

19. Pan A, Keum N, Okereke OI, Sun Q, Kivimaki M, Rubin RR, et al. Bidirectional association between depression and metabolic syndrome: A systematic review and meta-analysis of epidemiological studies. Diabetes Care. 2012 May;35(5):1171–80.

20. Luppino FS, De Wit LM, Bouvy PF, Stijnen T, Cuijpers P, Penninx BWJH, et al. Overweight, Obesity, and Depression A Systematic Review and Meta-analysis of Longitudinal Studies. Vol. 67, GEN PSYCHIATRY. 2010 Mar.

21. Fox CK, Gross AC, Rudser KD, Foy AMH, Kelly AS. Depression, Anxiety, and Severity of Obesity in Adolescents: Is Emotional Eating the Link? Clin Pediatr (Phila). 2016 Oct 1;55(12):1120–5.

22. Mühlig Y, Antel J, Föcker M, Hebebrand J. Are bidirectional associations of obesity and depression already apparent in childhood and adolescence as based on high-quality studies? A systematic review. Obesity Reviews. 2016 Mar 1;17(3):235–49.

23. Sutaria S, Devakumar D, Shikanai-Yasuda S, Das S, Saxena S. Is obesity associated with depression in children? Systematic review and meta-analysis. 2018;

24. Moradi M, Mozaffari H, Askari M, Azadbakht L. Association between overweight/obesity with depression, anxiety, low self-esteem, and body dissatisfaction in children and adolescents: a systematic review and meta-analysis of observational studies. Vol. 62, Critical Reviews in Food Science and Nutrition. Taylor and Francis Ltd.; 2021. p. 555–70.

25. Chaplin AB, Daniels NF, Ples D, Anderson RZ, Gregory-Jones A, Jones PB, et al. Longitudinal association between cardiovascular risk factors and depression in young people: A systematic review and meta-analysis of cohort studies. Psychol Med. 2023 Feb 25;53(3):1049–59.

26. Ma J, Rosas LG, Lv N, Xiao L, Snowden MB, Venditti EM, et al. Effect of Integrated Behavioral Weight Loss Treatment and Problem-Solving Therapy on Body Mass Index and Depressive Symptoms among Patients with Obesity and Depression: The RAINBOW Randomized Clinical Trial. JAMA - Journal of the American Medical Association. 2019 Mar 5;321(9):869–79.

27. Al-Khudairy L, Loveman E, Colquitt JL, Mead E, Johnson RE, Fraser H, et al. Diet, physical activity and behavioural interventions for the treatment of overweight or obese adolescents aged 12 to 17 years. Vol. 2017, Cochrane Database of Systematic Reviews. John Wiley and Sons Ltd; 2017.

28. Fuller NR, Burns J, Sainsbury A, Horsfield S, da Luz F, Zhang S, et al. Examining the association between depression and obesity during a weight management programme. Clin Obes. 2017 Dec 1;7(6):354–9.

29. Bai S, Zeledon LR, D’Amico EJ, Shoptaw S, Avina C, LaBorde AP, et al. Reducing health risk behaviors and improving depression in adolescents: A randomized controlled trial in primary care clinics. J Pediatr Psychol. 2018 Oct 1;43(9):1004–16.

30. Mansoor B, Rengasamy M, Hilton R, Porta G, He J, Spirito A, et al. The bidirectional relationship between body mass index and treatment outcome in adolescents with treatment-resistant depression. J Child Adolesc Psychopharmacol. 2013 Sep 1;23(7):458–67.

31. Tovilla-Zárate CA, Pérez-Mandujano A, Ramírez-González IR, Fresan A, Suarez-Mendez S, Martínez-Villaseñor E, et al. Vortioxetine versus sertraline in metabolic control, distress and depression in Mexican patients with type 2 diabetes. Ann Transl Med. 2019 Nov;7(22):656–656.

32. Leombruni P, Pierò A, Lavagnino L, Brustolin A, Campisi S, Fassino S. A randomized, double-blind trial comparing sertraline and fluoxetine 6-month treatment in obese patients with Binge Eating Disorder. Prog Neuropsychopharmacol Biol Psychiatry. 2008 Aug 1;32(6):1599–605.

